# An Automatic Computer-Based Method for Fast and Accurate Covid-19 Diagnosis

**DOI:** 10.1101/2020.07.02.20136721

**Authors:** Abdullah Al Jaid Jim, Ibrahim Rafi, Md. Sanaullah Chowdhury, Niloy Sikder, M. A. Parvez Mahmud, Saeed Rubaiee, Mehedi Masud, Anupam Kumar Bairagi, Kangkan Bhakta, Abdullah-Al Nahid

## Abstract

At present, the whole world is witnessing a horrifying outbreak caused by the Coronavirus Disease 2019 (COVID-19). The virus responsible for this disease is called SARS-CoV-2. It affects its victims’ respiratory system and causes severe lung inflammation, making it harder for them to breathe. The virus is airborne, and so has a high infection rate. Originated in China last December, the virus has spread across seven continents, affecting the population of over 210 countries, making it one of the fiercest pandemics ever recorded. Despite multiple independent and collaborative attempts to develop a vaccine or a cure, an effective solution is yet to come out. While the disease has put the world in a standstill, detecting the positive subjects and isolating them from the others as soon as possible is the only way to minimize its spread. However, many countries are currently experiencing a massive shortage of diagnostic equipment and medical personals. This insufficiency inspired us to work on a computer-based automatic method for the diagnosis of COVID-19. In this paper, we proposed a sequential Convolutional Neural Network (CNN)-based model to detect COVID-19 through analyzing Computed Tomography (CT) scan images. The model is capable of identifying the disease with almost 92.5% accuracy. We believe the implementation of this model will help the physicians and pathologists all over the world to single out the victims quickly and thus reduce the prevalence of COVID-19.

## 1. Introduction

Novel coronavirus disease (popularly known as COVID-19) is an outbreak in Wuhan, China, in late 2019 [1]. Severe acute respiratory syndrome coronavirus 2 (SARS-CoV-2) is responsible for this disease, whereas the World Health Organization (WHO) referred the virus as “the virus responsible for COVID-19” or “the COVID-19 virus” for general purposes [2]. SARS-CoV-2 virus has positive single stranded RNA (+ssRNA) having nucleocapsid and envelope at the outermost surface. Its genome structure is arranged in this +ssRNA, having a length of approximately 30 kilobases (kb) associated with a cap structure of 5’ and poly-A tail of 3’ [3]. This virus is one of the largest RNA viruses ever discovered. This COVID-19 in China was infecting hundreds of thousands of people and has become an epidemic. Soon this virus spread beyond the country border. Many COVID-19 cases have reported outside China quite quickly. This virus spreads from a host by droplet transmission or by airborne transmission [4]. Between these two transmissions, the first one infects from respiratory droplets coming out of a host, which insert into someone’s mouth and nose or conjunctiva (eyes). This process particularly occurs in near proximity (within 1 meter). The later one transmits from microbes within droplet nuclei. These nuclei are so small (<5µm) that they outlast in the air for a long duration. Anyone inhaling this air might have an infection. However, there is a similarity between these two transmissions. First, both require a host of COVID-19 to infect a new one, and second, the droplets must enter into a new body. Although the origin of the SARS-CoV-2 virus is still unexplored, we know how it spreads from one human being to another. Figure 1 illustrates a scenario of the transmission of this disease. At first, maybe one person got infected, who transmits that to the close ones. This condition less likely to be noticeable, and they continued infecting numerous people. Some were victims of close contact (from cough or sneeze), and some had the virus from the air or things that a COVID patient used (virus can survive up to a certain period on different materials). This chain remained unbroken, and soon this disease became an epidemic. When these infected people traveled to other parts of the world, they were the carrier for the parasite. Next, the locals got the COVID too, and again the mentioned chain began. Before we realized the circumstances, the epidemic turns into a global problem.

**Figure 1:**
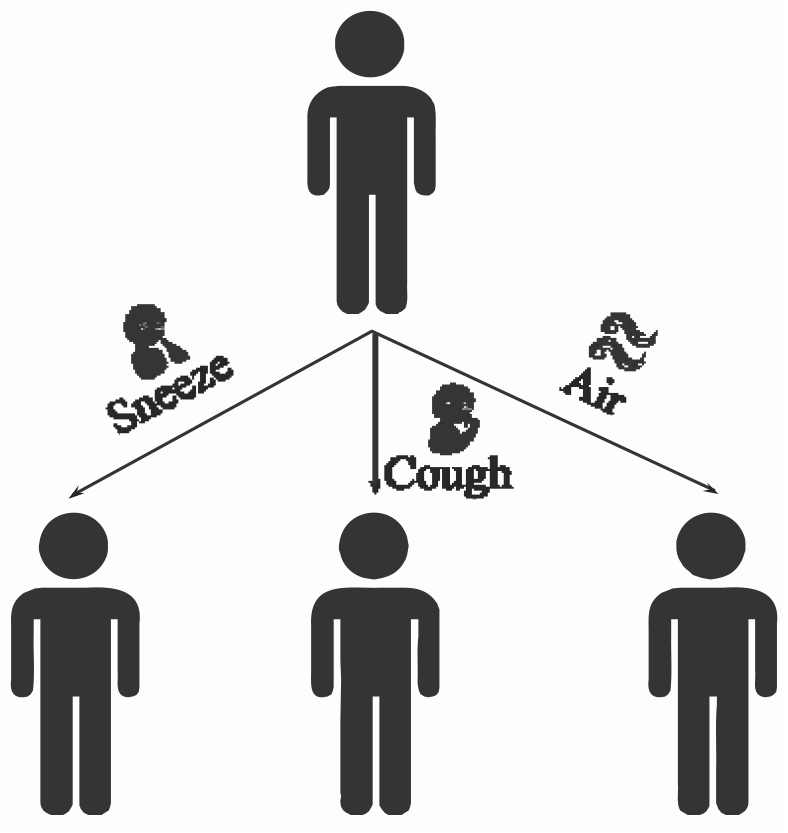
Transmission of the pandemic.

This SARS-CoV-2 virus transmits from human-to-human. Therefore, this spreads rapidly, and the World Health Organization announced COVID-19 a pandemic on March 12, 2020 [5]. It is threatening to whole humankind since it is taking lives as they are valueless. We have compared COVID-19 with some other viruses and mentioned the fatality rate in Table 1 [6]. According to the Worldometer, both Severe Acute Respiratory Syndrome (SARS) and Middle East Respiratory Syndrome (MERS) viruses have a higher death rate than COVID-19, in contrast, Swine flu has less death rate. However, the death rate is yet significant for people aging more than 50. The total number of COVID-19 hosts is doubled in 17 days across the globe, totaling around 2.16 million [7]. The number of COVID-19 cases increases at a geometric rate in almost every country.

**Table 1:** Fatality rate of different viruses

| Serial No. | Name of the virus | Fatality Rate |
| --- | --- | --- |
| 1 | Novel Coronavirus | 2% (estimated) |
| 2 | Swine Flu | 0.02% |
| 3 | SARS | 9.6% |
| 4 | MERS | 34% |

**Table 1:** Data Splitting Statistics

|  | Train | Validation | Test |
| --- | --- | --- | --- |
| **Non-COVID** | 146 | 15 | 34 |
| **COVID- 19** | 183 | 57 | 35 |
| **Total** | 329 | 73 | 69 |

So far, the number of COVID-19 patients increased very rapidly. Till May 2, 2020, the United States of America leads Spain, Italy, and the United Kingdom respectively in total COVID-19 cases. Whereas, total deaths are higher in the USA, Italy, UK, and Spain sequentially. In the USA, the place where most COVID-19 patients have found has the most recovered people. However, Belgium has the highest death rate (considering total deaths more than 1000). Further, Italy, France, the United Kingdom still have a high fatality rate. These pieces of information are summarized into the Table 2 according to [8].

**Table 2:**
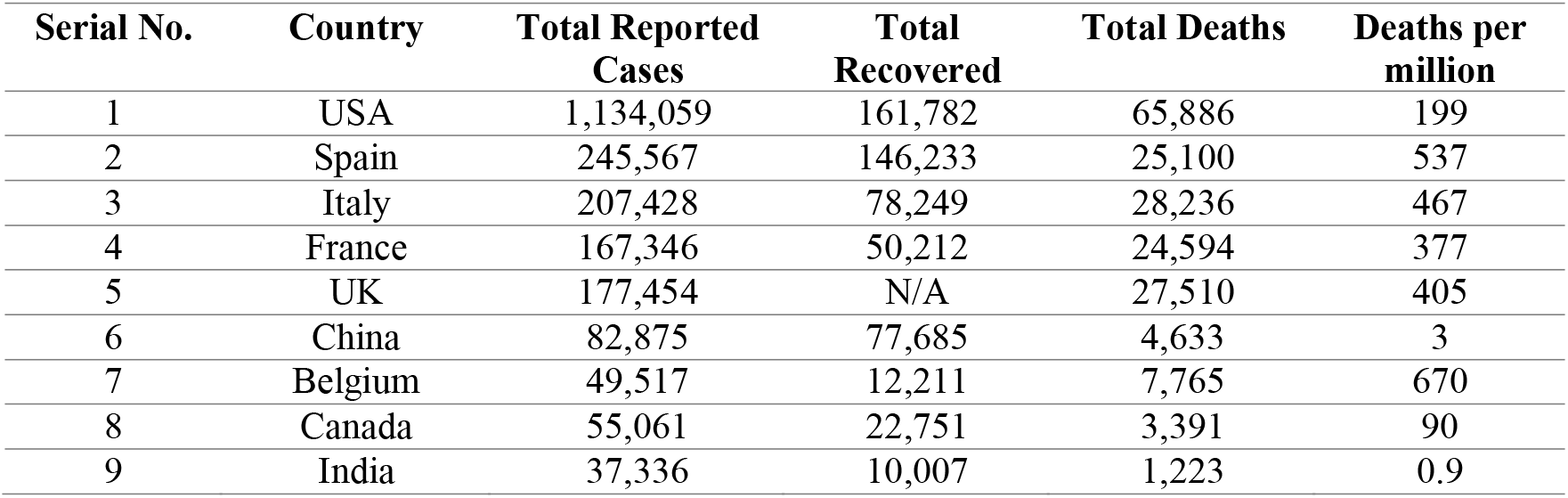
Fatality rate of COVID-19 in different countries

**Table 2:** Compare with the recent findings

| Work | Accuracy | Sensitivity | Specificity |
| --- | --- | --- | --- |
| [12] | 84.70% | 76.20% | N/A |
| [29] | 81.41% | 80.85% | 81.90% |
| Our Work | 92.48% | 94.17% | 89.58% |

Like every other disease, novel coronavirus disease has some symptoms too. Fever and sore throat are the very basic indications of this disease [9]. In most cases, pneumonia is identified in the patient around on the 9th day after infection [10]. Contrarily, pneumonia is caused by bacteria in general. There are three influential groups of pneumonia symptoms that are idiopathic interstitial pneumonia (IPF), nonspecific interstitial pneumonia (NSIP) and cryptogenic organizing pneumonia (COP) [11]. Detection of these categories requires bio-medical images such as x-ray images or CT (Computed Tomography) images to have a deep understanding of the stage and severity of pneumonia. These images are necessary for a doctor to prescribe any medicine and for the approach of treatment. However, we believe bacterial pneumonia and the one that COVID-19 patients have, might show different features in x-ray or CT images. Moreover, COVID-19 is quite a new disease on the face of the earth. Scientists are discovering something unique every day. There are a handful of skilled persons who can fight this pandemic. Training a person to contribute to this noble work is both time-consuming and troublesome. However, the solution can come from the machine learning perspective, such as Convolutional Neural Network (CNN). There is not enough information or features of COVID, which also motivates us to suggest a CNN based solution.

This model has first proposed by A.Krizhevsky et al. [13] in the year 2012 to classify images more accurately. It consisted of five convolutional layers along with max-pooling layers, three fully-connected layers, including a final “1000-way softmax”. To apply the computational power efficiently, they had adopted non-saturating neurons solely and also introduced “dropout” as the regularization method. Ever since then, this technique has made a renowned position in machine learning, especially in image classification (for example: [14], [15], [16], [17]) and image segmentation (for example: [18], [19], [20], [21]). Employing image classification in the medical sector is quite a recommended practice, for example, diabetic detection [22], cancer detection [23], pneumonia identification [24], [25]. A. A. Saraiva et al. [24] classified childhood pneumonia from x-ray images using CNN with an accuracy of 95.30%. I. Sirazitdinov et al. [28] also worked on pneumonia based on chest x-ray employing CNN. A CT image of the chest shows significant information about the COVID-19 disease. Inspired by the success of CNN architecture to identify and classify different diseases, researchers also give significant labor to classify and identify COVID-19 disease through the utilization of CNN architecture. [25] diagnosed COVID-19 pneumonia based on x-ray and CT images, where they have utilized a simple CNN model as well as a pre-trained AlexNet model. [31] proposed a light CNN model (basically a modified SqueezeNet CNN) to classify a set of CT images to classify COVID-19 disease. In their work, they obtained 83.00%, 85.00%, 81.00%, 81.73% of Accuracy, Sensitivity, Specificity, and Precision respectively. The work of [32] proposed a joined segmentation and classification approach based on a CNN model to diagnose COVID-19 disease. Lungs infection due to COVID-19 disease is identified through segmentation operation in the work [33], where the authors proposed Inf-Net and Semi-Inf-Net models. The authors of work [34] proposed a variation of the CNN model named as Detail-Oriented Capsule Networks (DECAPS) to classify the CT images for identification of COVID-19 diseases. To increase the amount of the training data, they utilized data augmentation operation based on the Conditional Generative Adversarial Network (GAN) structure. Along with the CT/x-ray images, ultrasound images also utilized for the COVID-19 disease identification. A lung ultrasound (POCUS) dataset is introduced in the work [35]. In this work, they utilized the POCOVID-Net model to detect automatically COVID-19 disease.

Getting inspired by these sorts of literature, we also performed CNN operation on CT scanned images in our work. The purpose of this research is to identify the pneumonia of COVID-19 patients by utilizing deep learning techniques specifically a CNN model. This may help mitigate the problem of skilled persons and also help to fight against Novel Coronavirus Disease. To do so, the rest of the paper is structured as follows. Section 2 provides a basic overview of how the work has been accomplished. Next, Section 3 is the representation of the dataset which we have adopted and somewhat modified for classification purpose. Some information concerning the dataset has also presented in this section. This work entirely depends on CNN. Therefore, Section 4 describes the application of CNN in our work. In Section 5, we have evaluated our model and made a comparison with recent findings. Finally, in Section 6, we have placed the concluding remarks and discussed possible future researches based on this work.

## 2. Methodology

This work utilizes a supervised classification model which classifies a set of images into, where represents Covid-19 and represents Non-COVID classes. COVID-19 and Non-COVID classes are the same as COVID-19 and Non-COVID ones mentioned in [12] respectively. After augmentation, the CT images from the dataset have split into training data (70% of data of the dataset) and testing data (which was 30% data) [12]. The supervised machine learning algorithm, in this case, is the CNN model hyper-tuned by the training dataset. Finally, the testing dataset has utilized to measure the performances of our model based on a few performance-measuring parameters. Figure 2 illustrates this overall working steps ostensibly.

**Figure 2:**
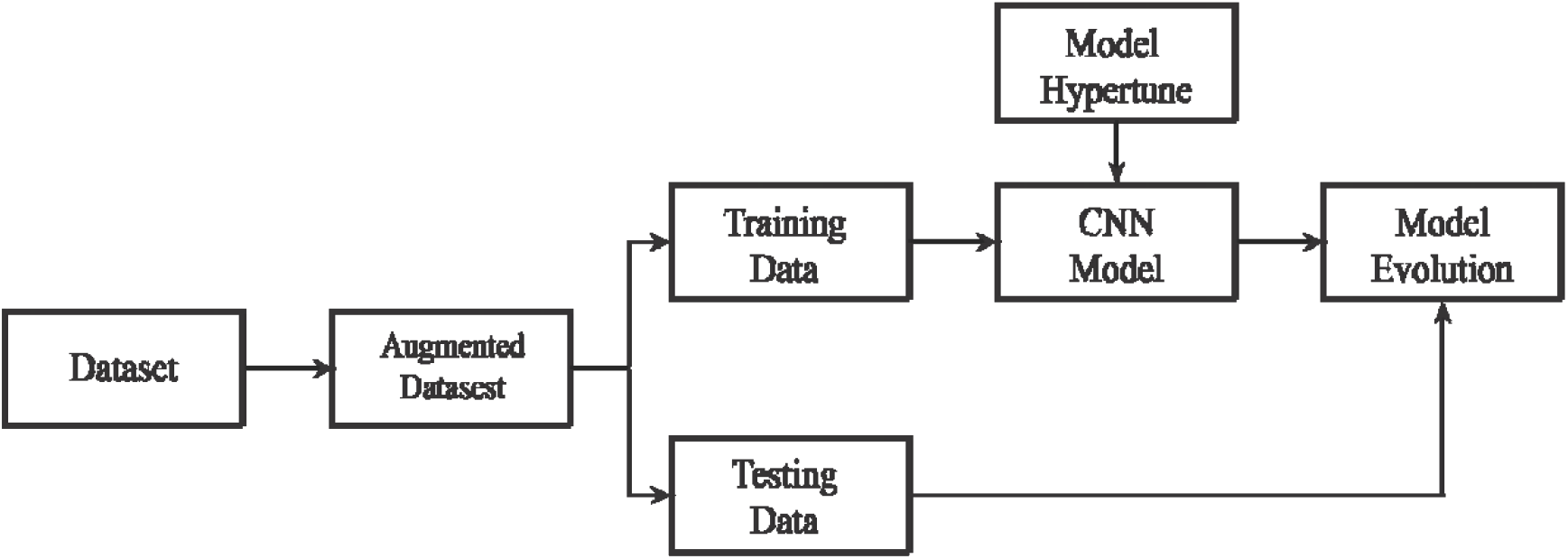
The total working steps of our research.

## 3. Dataset

For the prediction of pneumonia in COVID-19 patients, this work has utilized a dataset based on CT scan images [12]. For the creation of the dataset [12], the authors gathered 760 preprints about COVID-19. Finally, from those preprints, authors of [12] ended up with 275 CT scans and labeled them as COVID-19. The other cases where COVID-19 is not found they labeled them as Non-COVID. This dataset is accumulating more data over time, till April 28, 2020, they managed 349 CT images, from 216 patients of this disease. Figure 3a shows a CT scan imag of the respiratory system of COVID-19 patient, and Figure 3b shows the CT scan image of a non-COVID one. The dataset employed binary classification on CT images to result in COVID-19 or Non-COVID. A total of 195 images were negative for COVID-19, splitting the whole data into training, validation, and testing. The table 3 summaries this dataset.

**Figure 3:**
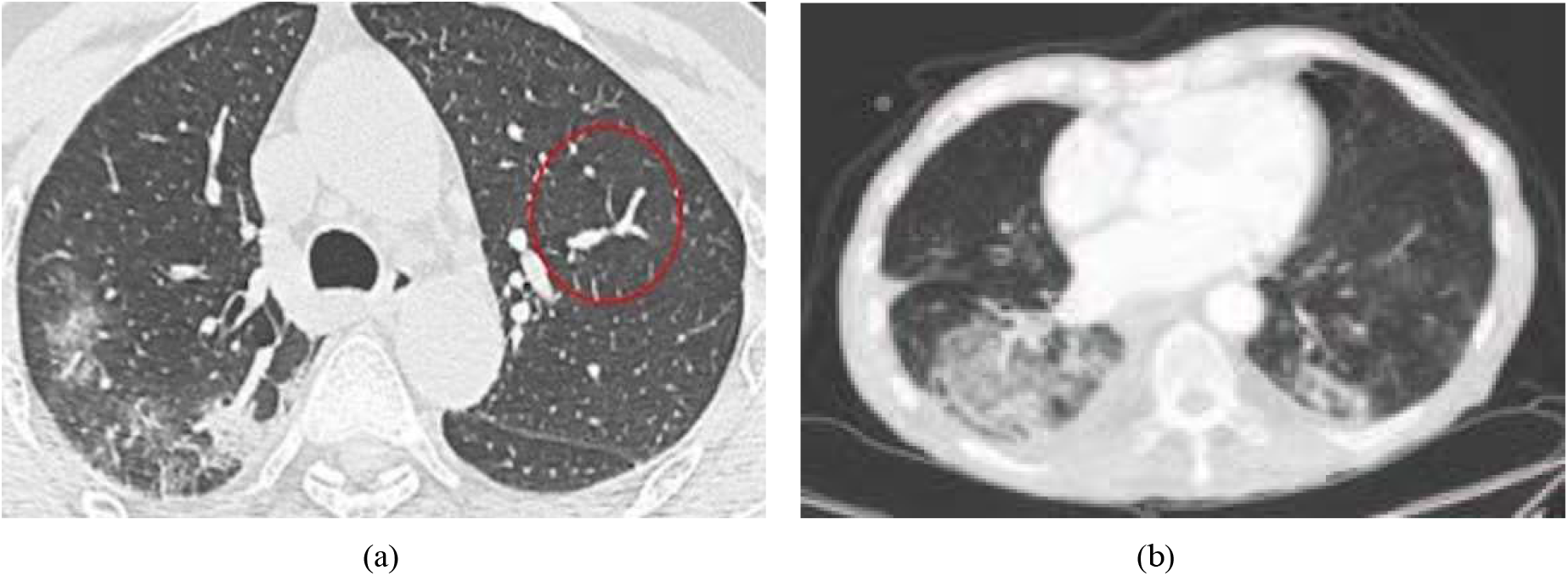
CT scan images of the respiratory system of (a) COVID-19 patient and (b) Non-COVID-19 person.

However, as the total amount of the images (471) is quite less to hyper-tune a CNN model efficiently, this work performs data augmentation operation such as (flip and rotation) on the dataset. These augmentation operations increase the original amount of images 14 times. Then the 70% of the images of the augmented dataset is utilized for training purposes and rest of the images for testing purpose.

## 4. Convolutional Neural Network

Learning is a distinct and spontaneous process for us humans, not for machines. Therefore, machines require extra assistance, various techniques, and methods. Convolutional Neural Network (CNN) is one of them, a computer-based deep learning technique that typically utilizes visuals. CNN is a biologically-inspired model to mimic the mammals’ visual activity [26]. It consists of two very fundamental operations - convolution & pooling. The operations are performed on different steps, and these steps often considered as layers in the model. There are some additional ones too, such as - an FC layer (Fully Connected layer) and the Softmax layer. Performing CNN on an image (of size 𝒥_*h*_ × 𝒥_*w*_) is implementing these operations in general. Convolution operation extracts features from an image like edge detection, color recognition, corner discovery and so on. This function may have filters (or kernel) of various numbers (𝒩_*f*_) with sizes (𝒩_*h*_ × 𝒩_*w*_), along with some other activities such as stride (𝒮) and padding (𝒫) to perform the complete procedure. After completing convolution, a featured matrix 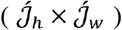 with lesser dimensions (or parameters) is obtained as output. The number of filters makes the number of images (𝒩_*f*_), further, height and width are calculated by the following equations.

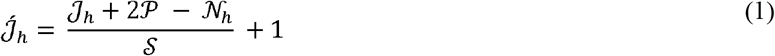

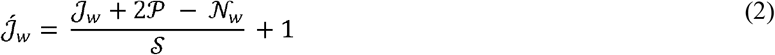

Pooling operation has similar properties like convolution by name filter size (𝓃_*h*_ × ×_*w*_), padding (𝒫_*p*_), and stride (𝒮_*p*_), performing the exact actions. Pooling solely reduces the computational power simply by decreasing the size of an image. An important information is this can reduce the height 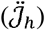 and the width 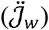 of the input keeping the 3^rd^ dimension (number of images, 𝒩_*f*_) unchanged. Further, they follow the same equations for conversion as convolution (equation 1 and 2).

The other layers (FC and softmax layers), which have been mentioned earlier, are hidden layers as they remain between the input and the output node. An image has as many nodes as the pixels, and output nodes are determined by the number of classes. In the FC layer, all the input nodes (*n*_out_) are flattened into a single column vector. Every node in this flattened layer has considered as a logits (𝒵_*i*_). These logits along with some other relations, calculate the softmax (𝓎_*i*_). The nodes (𝒳_*j*_), weights (𝒲_*ji*_), and biases (𝒷) combinedly calculate the logits, later these logits determine the softmax. Finally, the softmax produces C number of labels (*y*_*i*_), which are nothing but prediction with probability. All these properties are related and established by mathematical equations (equation 3 & 4).

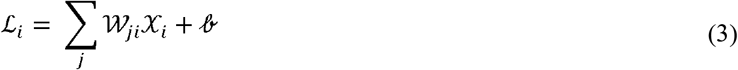

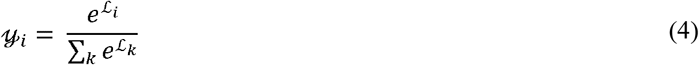

For prediction, we require a parameter that is a loss. There are three types of losses - *L*_1_ loss (ℒ_1_), *L*_2_ loss (ℒ_2_), and the cross-entropy loss (ℒ), all these have calculated comparing with original output (𝒴) with different approaches (represented in equation 5, 6 and 7). Among all the losses cross-entropy loss is widely accepted, and this is like a threshold value for the model.

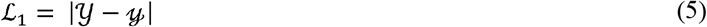

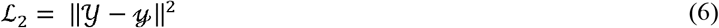

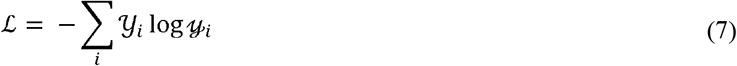

A CNN can be tuned to a certain loss to predict for this, both forward and backward propagation is crucial. At the initial condition, the model is not optimized. Therefore, it requires adjusting the weights (𝒲_*ji*_) and biases (𝒷) by determining the derivatives of loss w.r.t weight 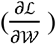, bias 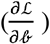, and logits 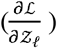. Backward propagation performs the work of adjusting the new values for learnable parameters (weights and biases), and forward propagation again calculates the loss. This process keeps repeating until obtaining the desired accuracy. After reaching the accuracy, the labels update with the probabilistic values. Backward propagation performs the work of adjusting the new values for learnable parameters (weights and biases), and forward propagation again calculates the loss. This process is called gradient descending, which keeps repeating until obtaining the desired accuracy. After reaching the accuracy, the labels update with the probabilistic values. The label having maximum value is the predicted output. The Figure 4 represents all the discussion above in a compact approach.

**Figure 4:**
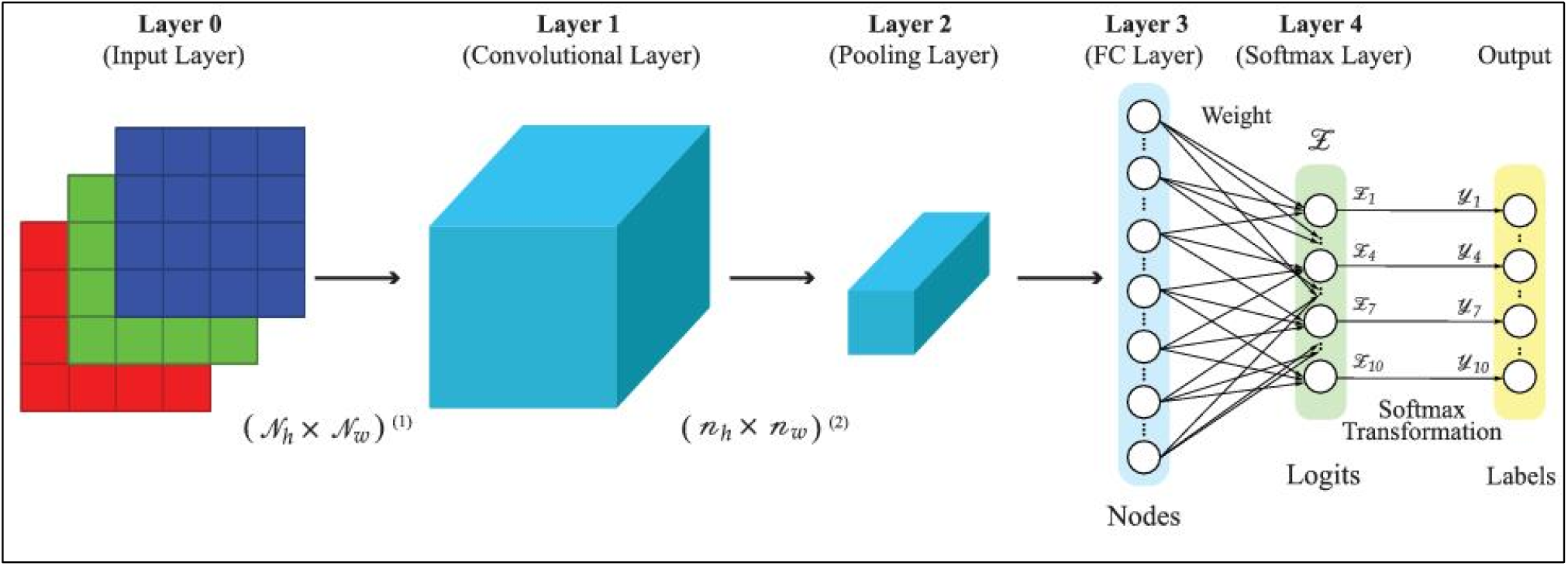
A basic model of Convolutional Neural Network.

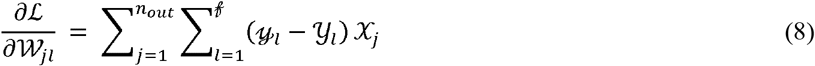

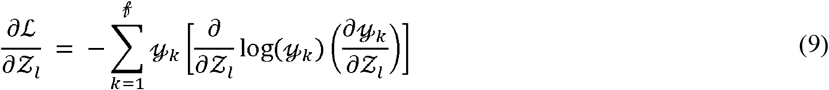

The sequential CNN model of our work is shown in Figure 5, where it takes an image as input. Then convolution is performed by a kernel of size 3 3 with the help of a function called Conv 2D. Next, the output of this function goes through an Activation function, more specifically, the Relu function. ReLU is the abbreviated form of the Rectified Linear Unit, which converts all the negative values of its input to zero. Max Pooling wit kernel size 2 2 will generate an output size of half of its input without changing any essential feature. This operation depends on taking the maximum values of the data, reducing the dimensions of an image. All these operations from convolution to pooling have done again to extract features. Further, all the neurons have flattened to a single column vector We have also introduced a 50% dropout to avoid the over fitting problem in the model. Now, these neurons have used to predict the classes in the Decision Layer with suitable relations. The process of making decisions and classifying them is called optimization.

**Figure 5:**
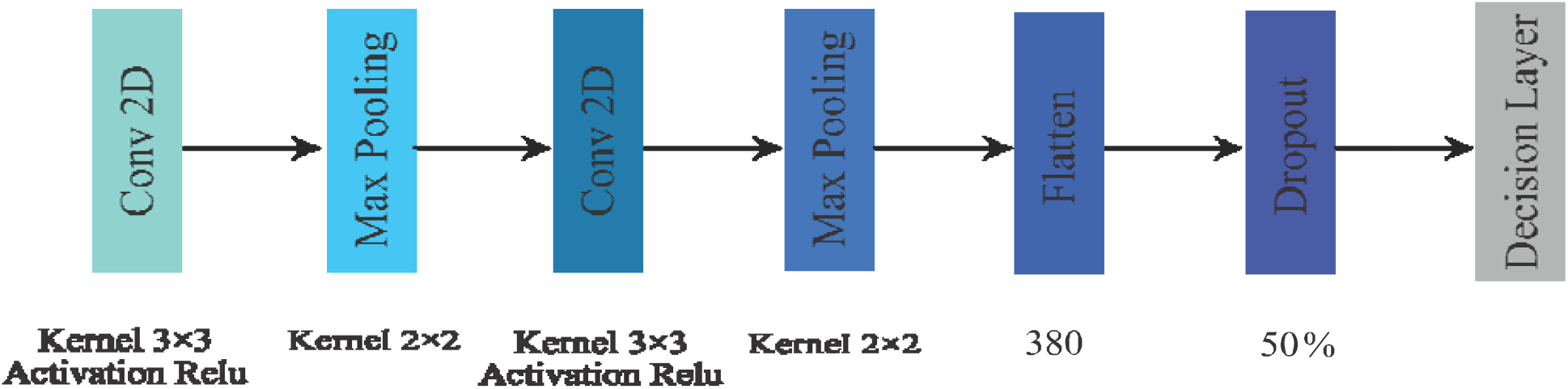
Schematic diagram of our proposed Convolutional Neural Network model.

## 5. Result and Discussion

### 5.1 Performance-Measuring Parameters

The major performance-measuring parameters for classifier algorithms are Accuracy, Recall or Sensitivity, Specificity, Precision, and F-measure [27]. Showing respect to this statement, we have calculated the mentioned parameters by using equation 10 to 14. We have also evaluated our findings based on them and compared them with other researches. All of them are greater, the better.

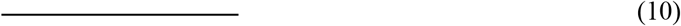

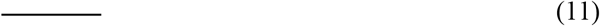

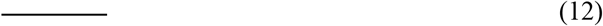

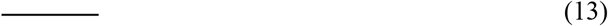

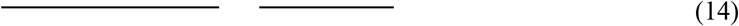

Here TP, TN, FN, and FP have their traditional meaning, True Positive, True Negative, False Negative, an False Positive respectively.

### 5.2 Description of the results

Accomplishing our work, we successfully classified two classes from the dataset. One is COVID-19 and, another one is Non-COVID. Our model is tuned for 200 epochs also we have calculated Accuracy, Recall, Specificity, Precision, and F-measure again based on test data, and recorded them for every epoch. Later, these data have used to plot the curves for Accuracy (Figure 6a), Sensitivity (Figure 6d), Precision (Figure 6c), and F1 score (Figure 7a).

**Figure 6:**
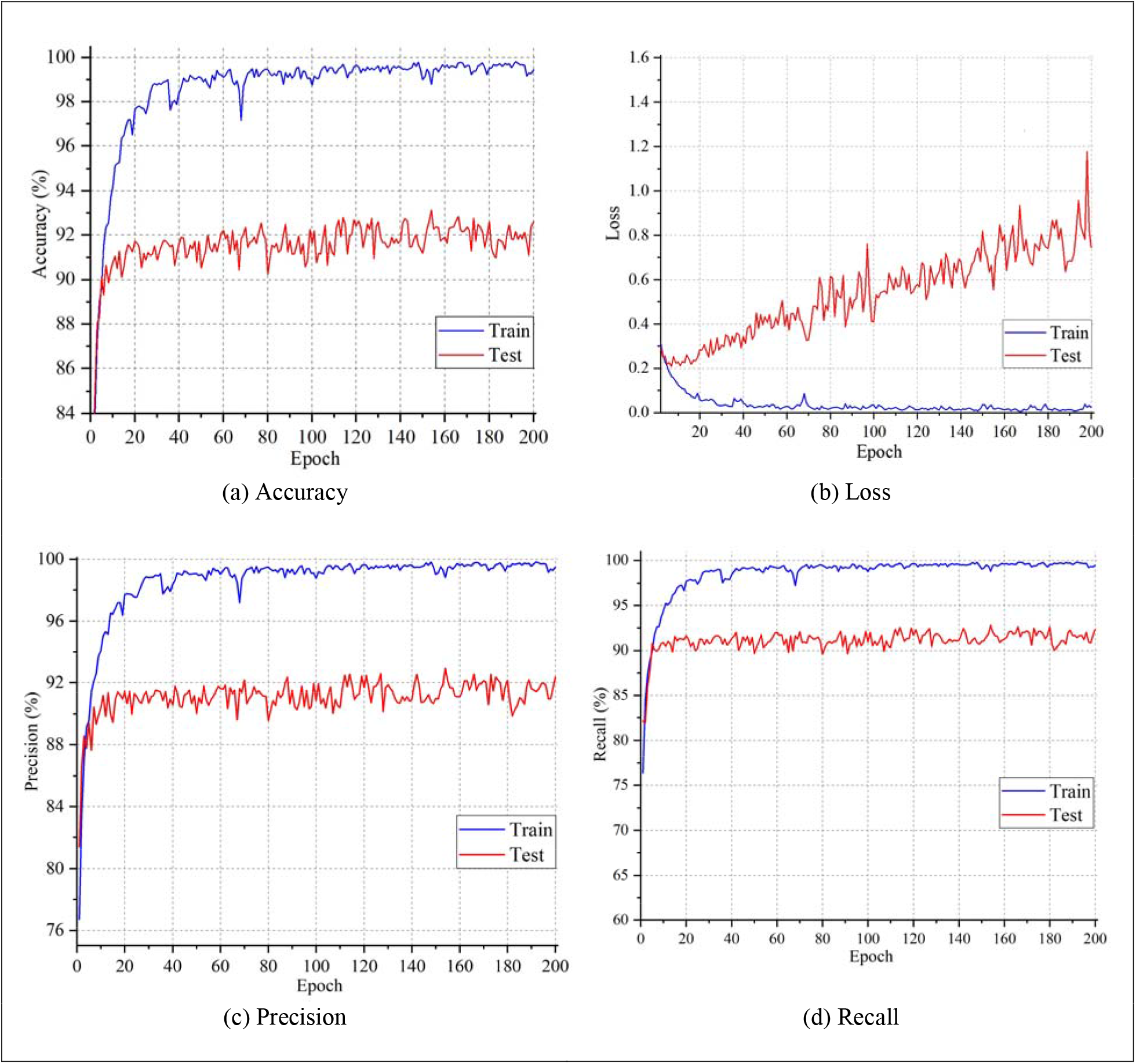
Different parameters for model evaluation.

**Figure 7:**
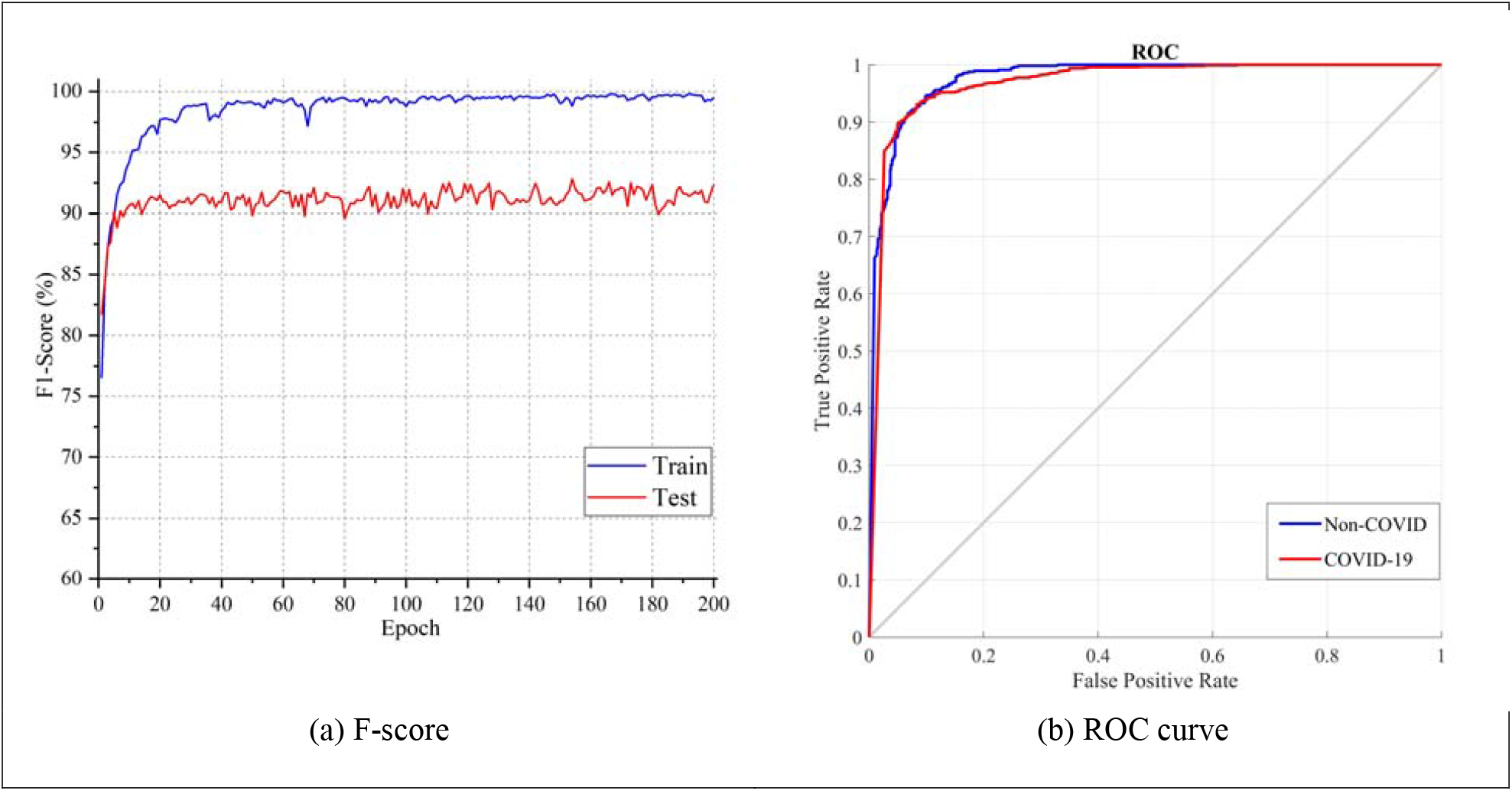
Other important parameters for the model.

The Accuracy, Sensitivity, Precision, and F1 score have measured for both training and testing data. If the parameter has calculated from training data, then it has a prefix as Training (e.g., Training-Accuracy) whereas, the one from testing data has a prefix as Testing (e.g., Testing-Precision). Figure 6 shows that Accuracy, Precision, and Sensitivity become more significant per epoch. This is an achievement for the model, as these parameters are better to be higher. In Figure 6a, initial accuracy was around 76% for training data and 82% for testing data. However, the values increase over every epoch gradually. Finally, on 200^th^ epoch, the value of Training-Accuracy reached almost to cent percent (99.46%) and Testing-Accuracy to 92.61%. The value of Testing-Accuracy is rather satisfactory. For every epoch, we have calculated the loss and saved the values. These values are used to plot the curve of loss in Figure 6b. The loss calculated on the training dataset is comparatively low comparing to the testing dataset valuing almost zero. Each epoch increases accuracy and decreases the loss of training data. Figure 6c shows somewhat similar characteristics of precision as accuracy. Initially, Testing-Precision was greater than that of the Training one, but they slowly increased and Training-Precision became bigger. The final values of Training-Precision and Testing-Precision are 99.48% and 92.36% sequentially. Figure 6d illustrates an upward rising sensitivity curve. Training-Sensitivity and Testing-Sensitivity both moved to 90% in just around 6 epochs, making final values 99.44% and 92.37% respectively. The F1 score was so low at the beginning, as shown in Figure 7a. Both the F1 score started with significant values (more than 75%) and managed to 99.46% and 92.36% at the end of 200 epochs. The Receiver Operating Characteristic curve (ROC curve) is a vital parameter to determine the diagnostic ability of a binary classification CNN model graphically [30]. This curve is plotted based on True Positive Rate (TPR) or sensitivity against False Positive Rate (FPR). Figure 7b illustrates the ROC curve of our model.

Figure 8 is the graphical representation of the confusion matrix where 920, 507, 59, and 57 values exhibit the values of TP, TN, FP, and FN sequentially. Here, TP, TN, FP, and FN bear their conventional definition i.e., True Positive, True Negative, False Positive, and False Negative respectively. These values are essential for determining Accuracy, Recall, Specificity, Precision, and F-measure. From the confusion matrix, the Accuracy, Sensitivity, Specificity, Precision, and F1 score of our model are 92.48%, 94.17%, 89.58%, 93.97%, and 94.07% respectively (by applying equation 10 to 14). Lastly, from sensitivity and specificity, the calculated balanced accuracy is 91.88%.

**Figure 8:**
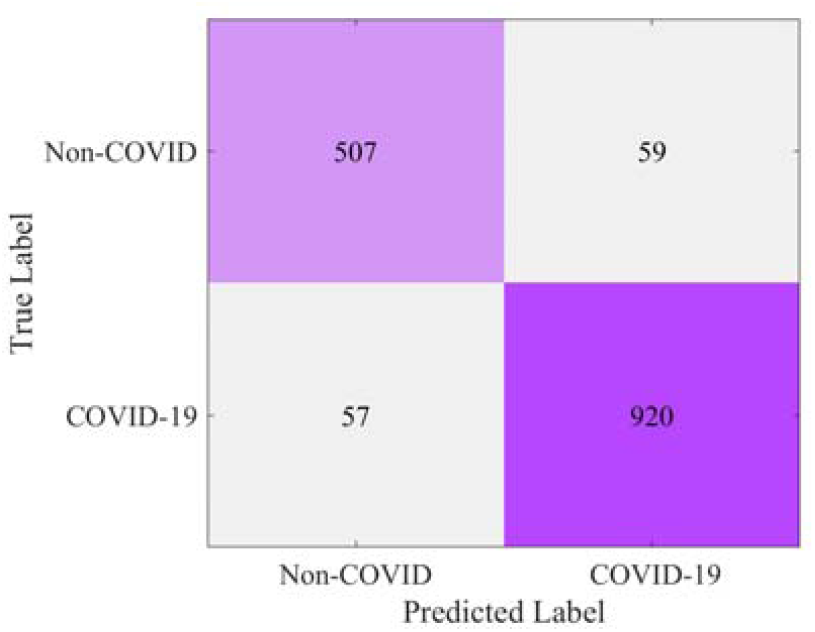
Confusion Matrix of the Model.

We have compared our findings with others and presented in Table 4. For comparison, we were enthused CNN model based work. Our work has better accuracy as well as sensitivity than both [12] and [29]. Our model has 7.78% and 11.07% more accuracy than [12] and [29] individually. This model also assures better sensitivity than [12] and [29], that is approximately 17.97% and 13.32% respectively. It has greater specificity and balanced accuracy than [29]. Comparing these parameters, we can claim that our model is more suitable for classifying (COVID-19 or Non-COVID) CT scan images than these two models.

## 6. Conclusion

This paper described a novel COVID-19 diagnosis method in the form of a binary classification task using a sequential CNN, based on the knowledge attained from CT scan images. The acquired results suggest that the model is highly potent in carrying out its job and can reach a maximum accuracy of 92.48%. Other associated parameters verify the authenticity of the result obtained by the proposed method and assert its superiority over the previous ones. The sole purpose of this study is to assist the global fight against COVID-19 that the medical professionals are fighting relentlessly by providing them an easy and effective means of diagnosis of the disease. Being academics, we hope this model will be implemented and used worldwide, and play a part in putting this pandemic behind us.

## Data Availability

The data set is available in git-hub repository.
It is free to use for all.

## References

[1] Novel, Coronavirus Pneumonia Emergency Response Epidemiology. “The epidemiological characteristics of an outbreak of 2019 novel coronavirus diseases (COVID-19) in China.” Zhonghua liu xing bing xue za zhi= Zhonghua liuxingbingxue zazhi 41.2 (2020): 145.

[2] World Health Organization. Available online: https://www.who.int/ emergencies/diseases/novel-coronavirus-2019/technical-guidance/ naming-the-coronavirus-disease-(covid-2019)-and-the-virus-that-causes-it (accessed on 2 May 2020)

[3] Cascella, M., Rajnik, M., Cuomo, A., Dulebohn, S. C., & Di Napoli, R. (2020). Features, evaluation and treatment coronavirus (COVID-19). In StatPearls [Internet]. StatPearls Publishing.

[4] Singhal, T. (2020). A review of coronavirus disease-2019 (COVID-19). The Indian Journal of Pediatrics, 1–6.

[5] World Health Organization. Available online: http://www.euro.who.int/en/ health-topics/health-emergencies/coronavirus-covid-19/news/news/2020/3/ who-announces-covid-19-outbreak-a-pandemic (accessed on 2 May 2020)

[6] Worldometer. Available online: https://www.worldometers.info/coronavirus/coronavirus-death-rate/ (accessed on 2 May 2020).

[7] Roser, M., Ritchie, H., & Ortiz-Ospina, E. (2020). Coronavirus Disease (COVID-19)–Statistics and Research. Our World in Data.

[8] Worldometer. Available online: https://www.worldometers.info/coronavirus/ (accessed on 2 May 2020)

[9] Day, M. (2020). Covid-19: ibuprofen should not be used for managing symptoms, say doctors and scientists.

[10] Holshue, M. L., DeBolt, C., Lindquist, S., Lofy, K. H., Wiesman, J., Bruce, H., & Diaz, G. (2020). First case of 2019 novel coronavirus in the United States. New England Journal of Medicine.

[11] Tachibana, R., Kido, S., Tanaka, N., & Matsumoto, T. (2004, June). Power spectral analysis of idiopathic interstitial pneumonias in high resolution CT images. In International Congress Series (Vol. 1268, pp. 961–966). Elsevier.

[12] Zhao, J., Zhang, Y., He, X., & Xie, P. (2020). Covid-ct-dataset: A ct scan dataset about covid-19. arXiv preprint 2003.13865.

[13] Krizhevsky, A., Sutskever, I., & Hinton, G. E. (2012). Imagenet classification with deep convolutional neural networks. In Advances in neural information processing systems (pp. 1097–1105).

[14] Wei, Y., Xia, W., Lin, M., Huang, J., Ni, B., Dong, J., & Yan, S. (2015). HCP: A flexible CNN framework for multi-label image classification. IEEE transactions on pattern analysis and machine intelligence, 38(9), 1901–1907.

[15] Hou, L., Samaras, D., Kurc, T. M., Gao, Y., Davis, J. E., & Saltz, J. H. (2016). Patch-based convolutional neural network for whole slide tissue image classification. In Proceedings of the IEEE conference on computer vision and pattern recognition (pp. 2424–2433).

[16] Akata, Z., Reed, S., Walter, D., Lee, H., & Schiele, B. (2015). Evaluation of output embeddings for fine-grained image classification. In Proceedings of the IEEE Conference on Computer Vision and Pattern Recognition (pp. 2927–2936).

[17] Liu, B., Yu, X., Zhang, P., Yu, A., Fu, Q., & Wei, X. (2017). Supervised deep feature extraction for hyperspectral image classification. IEEE Transactions on Geoscience and Remote Sensing, 56(4), 1909–1921.

[18] Liu, F., Lin, G., & Shen, C. (2015). CRF learning with CNN features for image segmentation. Pattern Recognition, 48(10), 2983–2992.

[19] Vilariño, D. L., Brea, V. M., Cabello, D., & Pardo, J. M. (1998). Discrete-time CNN for image segmentation by active contours. Pattern Recognition Letters, 19(8), 721–734.

[20] A Bao, S., & Chung, A. C. (2018). Multi-scale structured CNN with label consistency for brain MR image segmentation. Computer Methods in Biomechanics and Biomedical Engineering: Imaging & Visualization, 6(1), 113–117.

[21] Liu, Z., Li, X., Luo, P., Loy, C. C., & Tang, X. (2015). Semantic image segmentation via deep parsing network. In Proceedings of the IEEE international conference on computer vision (pp. 1377–1385).

[22] ASwapna, G., Kp, S., & Vinayakumar, R. (2018). Automated detection of diabetes using CNN and CNN-LSTM network and heart rate signals. Procedia computer science, 132, 1253–1262.

[23] Tan, Y. J., Sim, K. S., & Ting, F. F. (2017, November). Breast cancer detection using convolutional neural networks for mammogram imaging system. In 2017 International Conference on Robotics, Automation and Sciences (ICORAS) (pp. 1-5). IEEE.

[24] Saraiva, A., Ferreira, N., Sousa, L., Carvalho da Costa, N., Sousa, J., Santos, D., & Soares, S. (2019). Classification of Images of Childhood Pneumonia using Convolutional Neural Networks. In 6th International Conference on Bioimaging (pp. 112–119).

[25] Maghdid, H. S., Asaad, A. T., Ghafoor, K. Z., Sadiq, A. S., & Khan, M. K. (2020). Diagnosing COVID-19 pneumonia from X-ray and CT images using deep learning and transfer learning algorithms. arXiv preprint 2004.00038.

[26] Hubel, D. H., & Wiesel, T. N. (1965). Receptive fields and functional architecture in two nonstriate visual areas (18 and 19) of the cat. Journal of neurophysiology, 28(2), 229–289.

[27] Goutte, C.; Gaussier, E. A Probabilistic Interpretation of Precision, Recall and F-Score, with Implication for Evaluation. In; 2010.

[28] Sirazitdinov, I., Kholiavchenko, M., Mustafaev, T., Yixuan, Y., Kuleev, R., & Ibragimov, B. (2019). Deep neural network ensemble for pneumonia localization from a large-scale chest x-ray database. Computers & Electrical Engineering, 78, 388–399.

[29] Loey, M., Smarandache, F., & Khalifa, N. E. M. (2020). A Deep Transfer Learning Model with Classical Data Augmentation and CGAN to Detect COVID-19 from Chest CT.

[30] Flach, P. A. (2016). ROC analysis. In Encyclopedia of Machine Learning and Data Mining (pp. 1–8). Springer.

[31] Polsinelli, M., Cinque, L., & Placidi, G. (2020). A Light CNN for detecting COVID-19 from CT scans of the chest.

[32] Wu, Y., Gao, S., Mei, J., Xu, J., Fan, D., Zhao, C., & Cheng, M. (2020). JCS: An Explainable COVID-19 Diagnosis System by Joint Classification and Segmentation. ArXiv, abs/2004.07054.

[33] Fan, D., Zhou, T., Ji, G., Zhou, Y., Chen, G., Fu, H., Shen, J., & Shao, L. (2020). Inf-Net: Automatic COVID-19 Lung Infection Segmentation from CT Scans. medRxiv.

[34] Mobiny, A., Cicalese, P.A., Zare, S., Yuan, P., Abavisani, M., Wu, C.C., Ahuja, J., Groot, P.M., & Nguyen, H.V. (2020). Radiologist-Level COVID-19 Detection Using CT Scans with Detail-Oriented Capsule Networks. ArXiv, abs/2004.07407.

[35] Born, J., Brandle, G., Cossio, M., Disdier, M., Goulet, J., Roulin, J., & Wiedemann, N. (2020). POCOVID-Net: Automatic Detection of COVID-19 from a New Lung Ultrasound Imaging Dataset (POCUS).

